# Assessment of the Knowledge, Attitude, and Practice of Students toward Passive Smoking at the University of Abuja, Nigeria

**DOI:** 10.1101/2024.11.24.24317480

**Authors:** George Killian Jethniel, Stephen Ikechukwu Azumara, Iliya Ishaya, Philip Tobi Akpobasa, Mercy Onyeani Atu

**Affiliations:** Department of Community Medicine, College of Health Sciences, University of Abuja, Nigeria; Department of Clinical Pharmacy and Pharmacy Management, University of Nigeria, Nsukka, Enugu State, Nigeria; Department of Community Medicine, College of Medical Sciences, Ahmadu Bello University, Zaria, Nigeria; Department of Clinical Pharmacy and Pharmacy Administration, University of Ibadan, Oyo State. Nigeria

**Keywords:** Passive smoke, students, health effects, smoke laws, Nigeria

## Abstract

**Background:** Passive smoking, also known as involuntary or secondhand smoking, is a substantial public severe health concern, leading to illness, disability, and death. The attitude, practice, and knowledge of students towards passive smoking have not been adequately addressed in Nigeria. Therefore, this study aimed to investigate the attitude, practice, and knowledge of passive smoking among undergraduate students at the University of Abuja.

**Methods:** This cross-sectional study employed a multi-stage sampling technique to include 355 students at the University of Abuja, Nigeria. A self-completed questionnaire was utilized to gather information on socio-demographics, attitudes regarding passive smoking, practice of passive smoke exposure, and awareness of passive smoking.

**Results:** Exposure to passive smoke was reported by 60.4% of the participants, 80.6% exhibited a solid comprehension of passive smoking, and most of the participants (98.9%) had a positive attitude towards passive smoking. Characteristics such as level of study and age were identified as statistically significant, showing p-values of 0.000 and 0.001, respectively.

**Conclusion:** Our study highlights that even though students at the University of Abuja exhibit a commendable level of knowledge and a positive attitude toward passive smoking prevention, the high prevalence of exposure indicates a significant gap between awareness and practice. Therefore, university authorities in Nigeria should develop educational initiatives to decrease exposure to passive smoke and establish control policies towards mitigating exposure to passive smoking among students in Nigerian universities.

## INTRODUCTION

Smoking involves inhaling the smoke produced by burning tobacco found in cigars, cigarettes, and pipes. It is the leading cause of preventable illness and early mortality worldwide, with around 1.3 billion individuals engaging in the habit, predominantly in developing nations, where roughly 80% of smokers reside.^1^ Previous studies on tobacco use in Nigeria indicated that the smoking prevalence was 31.9% in certain urban areas and 17.6% in selected rural regions. ^2^ A recent survey conducted in Osogbo, Osun State, found that 8.7% of participants were current smokers. ^3^ Additionally, It is estimated that approximately 207,000 children in the United Kingdom take up smoking each year. ^4^

Passive smoking, also known as involuntary or secondhand smoking, occurs when someone breathes in smoke produced by burning tobacco products or exhaled by someone who smokes. Annually, 600,000 morbidities are recorded as a result of exposure of non-smokers to passive smoking globally.^5^ This exposure is estimated to have led to approximately 21,400 deaths from lung cancer, 165,000 from lower respiratory infections, 36,900 from asthma, and 379,000 from ischemic heart disease.^6^ In 2004, worldwide, around 40% of children, 33% of male non-smokers and 35% of female non-smokers were involved in secondhand smoke.^5^ The diseases that lead to death as a result of passive smoking include respiratory infections and asthma in children, as well as conditions like ischemic heart disease, lung cancer, and asthma, amounting to a disease burden equivalent to 10.9 million years of life adjusted for disability (DALYs) according to WHO.^7^ A complex mixture of chemicals is generated from the burning and smoking of tobacco, and these chemicals result in some adverse health outcomes, including respiratory and cardiovascular diseases and some cancer types.^8^ Moreover, they can worsen preexisting conditions like asthma.^8^ The smoke that emanates from a burning cigarette is termed “sidestream smoke,” whereas exhaled smoke is called exhaled mainstream smoke, and both of them are collectively often termed environmental tobacco smoke (ETS) or secondhand smoke (SHS).^9^ When the residues of secondhand smoke that stick to indoor dust and surfaces or just lying around are reemitted into the air, it is referred to as thirdhand smoke.^10^ Passive smoking comprises both secondhand smoking (SHS) and thirdhand smoking (THS). Friends and families of smokers are exposed to passive smoking. Furthermore, children and pregnant women are at risk of developing adverse health conditions from passive smoking.^8^ Globally, people are affected by the harmful effects of smoking either directly by smoking themselves or indirectly by being exposed to it as others smoke. Therefore, it is crucial to emphasize the prevention of passive smoking because people exposed might not know the harmful effects or might not be able to control the air pollution.

Tobacco stands as the primary preventable cause of death across the globe.^11^ Cigarette smoking has emerged as a significant public health issue more than at any other time in history.^12^ The World Health Organization (WHO) estimates that approximately 1.3 billion individuals worldwide use tobacco products, with 80% of these individuals residing in countries with low and middle incomes, which are frequently targeted by tobacco marketing and interference.^8^ Consequently, a great deal of tobacco products are consumed in developing countries, making air pollution with tobacco products a major concern. There are common misconceptions about passive smoking. Quite a number of people believe that while smoking bothers other people, it is not dangerous, poses no health hazards to other people, and does not affect one’s children.^13^ The United States Centre for Disease Control (CDC) has debunked these wrong ideas.^13^

Tobacco smoke comprises over 7,000 chemical substances and compounds, which are either vapour or solids,^14^ with at least 70 of these substances identified as carcinogenic.^15^ These substances include nicotine, polycyclic aromatic hydrocarbons, nitrated PAH, benzo[a]pyrene, thiocyanate, nitrolactones, and chrysene.^14^ The danger of smoking and inhaling cigarette smoke stems from the compounds in the cigarette, some in the tobacco, and some due to combustion.^15^ All cigarettes are dangerous and have no safe amounts, and exposure to them can cause both immediate and long-term consequences.^15^

University students are easily exposed to passive smoke because the school environment comprises varying populations, including friends, acquaintances, and teaching and non-teaching staff. Students face several challenges during their stay at the university, and it is not uncommon for them to turn to smoking and exposing other students to passive smoking. Some students also smoke in the hostels, classrooms, and gatherings, putting other non-smokers at risk. The early years at the university mark the initiation period of smoking.^11^ These students have a higher risk of future smoking than their age mates not attending universities.^16^ To attract university students to smoke, the tobacco industry targets explicitly their promotions at bars, clubs, and beer parlours close to university campuses, and most of these universities do not have a comprehensive ban on smoking.^16^ The likelihood of smoking among university students is heavily influenced by unsafe behaviors, including marijuana use, having multiple sexual partners, alcoholism, cultism, poor academic performance, stress, and depression.^17^ These factors expose other students to passive smoking from students who smoke because they hang out with students involved in such activities in classrooms, hostels, and other places on campus.

Smoking is addressed by the WHO Framework Convention on Tobacco Control (WHO FCTC), a treaty grounded in evidence that emphasizes everyone’s entitlement to optimal health. This treaty was created as a response to the global tobacco crisis.^18^ Nigeria officially joined the WHO FCTC on January 18, 2006.^19^ Some of the regulations surrounding tobacco use include the prohibition of smoking in areas free from smoke, prohibiting tobacco advertising, sponsorships, and promotions, the requirement of text-only warnings on tobacco packaging and labelling, the regulation and disclosure of cigarette contents, and restrictions on sales (such as prohibiting the sale of single cigarettes, small packs, and internet sales).^18,19^

All over the world, enhancing knowledge, practices, and attitudes is regarded as a key strategy for minimizing exposure to passive smoking.^20^ Low index of knowledge, practice, and attitude of passive smoking is observed among young people,^21^ who make up a bulk of the university students’ population. University students are exposed to passive smoking because of their environment, which increases their chances of coming close to persons who smoke. Little is known about passive smoking and associated health hazards among university students in the northern region of Nigeria and the University of Abuja. Hence, this study aimed to assess the attitude, practice, and knowledge of passive smoking among undergraduate students at the University of Abuja.

## METHODS

### Study design

The study used a descriptive cross-sectional design. Planned dates for data collection were over the period of approximately 2 months, from 2021-06-1 to 2021-07-30. The first reported response was recorded on 2021-06-1, while the last recorded response was on 2021-07-30.

### Ethical Approval

All participants were adults and provided informed consent before participation. An information sheet detailing the study objectives and participants’ rights was provided. Ethics approval was obtained from the committee responsible for health research ethics at the University of Abuja Teaching Hospital, and permission was also granted by the University’s Students’ Affairs Division.

### Study setting

The study took place at the University of Abuja, located in Gwagwalada. Gwagwalada is one of the six local government areas of the Federal Capital Territory (FCT), Nigeria. The University of Abuja, established on January 1, 1988, following the amendments made to Decree No. 110 of 1992, is a public tertiary institution offering conventional and distance learning programs.

### Study population and sampling method

The study population consists of regular undergraduate students enrolled in academic programs at the University of Abuja. In total, partial or complete responses were captured from 341 individuals. The flowchart for participation in the study is shown in Figure 1. The study employed a multistage sampling method. In the initial phase, a stratified sampling technique was used to group the colleges/faculties in the University into eleven (11) strata. In the second stage, simple random sampling was utilized by way of balloting without replacement to select eight (8) colleges/faculties out of the eleven. The third stage involved a random sampling of approximately the same number of students from the selected colleges/faculties.

**Figure 1:**
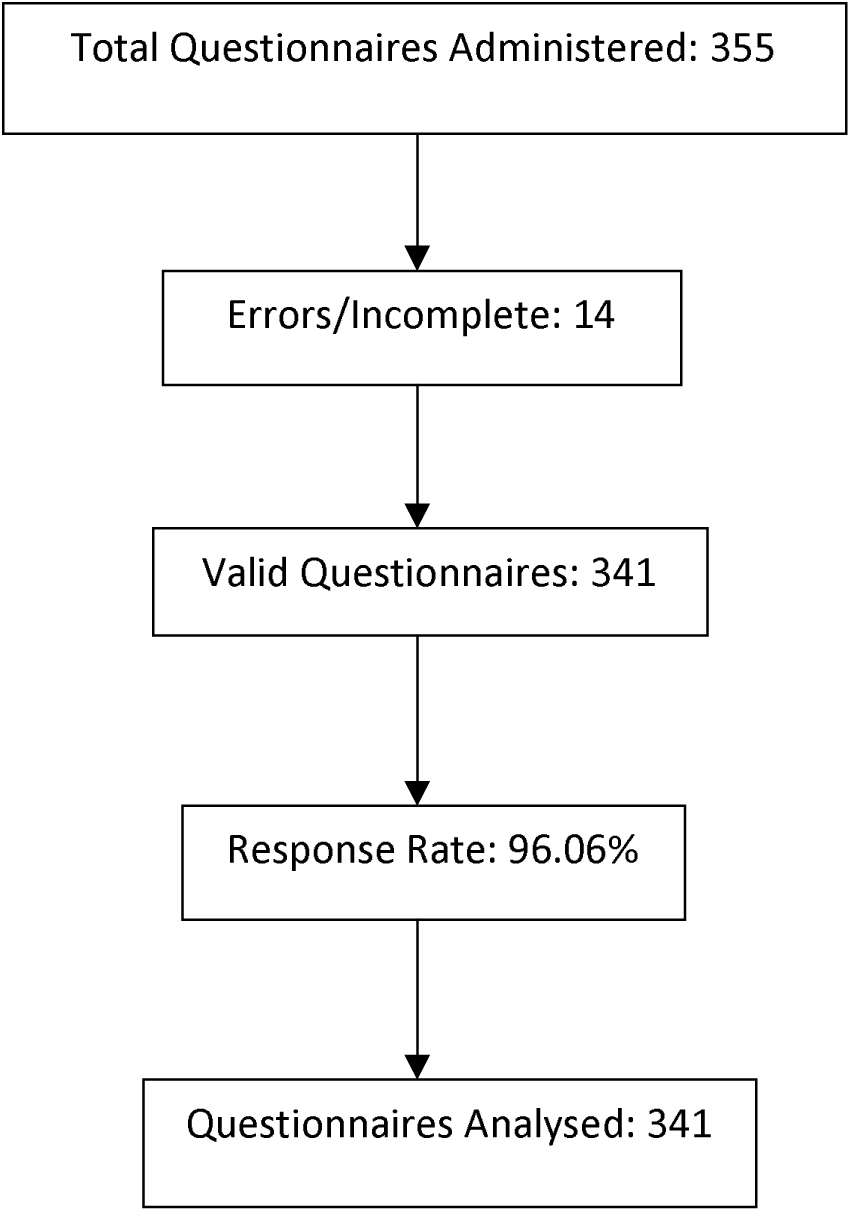
Flowchart of Study Participation and Response Analysis

### Inclusion criteria

Regular undergraduate students enrolled in academic programs at the University of Abuja.

### Exclusion criteria

Non-regular undergraduate students and all postgraduate students enrolled in academic programs at the University of Abuja were excluded.

### Sample Size Calculation

The sample size was determined using the Leslie-Kish formula:

**n**^0^**=Z^2^pq/d**^2^, Where;
n^0^ = the necessary sample size overall
Z = the Z-score associated with the specified confidence level (1.96 is the Z-score for a 95% confidence level)
p = the point prevalence of passive smoking in undergraduate students from previous studies and was taken to be 69.9% (0.70).
q = 1 - p represents the probability of the outcome not occurring.
d = is the margin of error set at 0.05
n0 = [(1.96)^2 * 0.70 * (1 - 0.70)] / (0.05)^2 = 322.56 ≈ 323.

Taking into account a 10% non-response rate, the final sample size was adjusted to 355 participants.

### Data collection tool

Information was collected through a semi-structured, pre-tested, self-administered questionnaire comprising four sections: Section A: Collected sociodemographic information. Section B: Assessed participants’ knowledge of passive smoking through 9 questions. A total score of 9 was possible, with scores of 5 or higher considered indicative of good knowledge. Section C: Measured students’ attitudes toward passive smoking using a 7-item, 5-point Likert scale. Scores of 18 or higher indicated a positive attitude. Section D: Evaluated the practice of passive smoking among respondents.

### Data Management and Analysis

Analysis of the data was performed using IBM SPSS version 23.0 (IBM Corp., Armonk, NY, USA). Categorical variables were summarized through frequencies and percentages, while continuous variables were reported as means along with standard deviations. A p-value of less than 0.05 was regarded as statistically significant, corresponding to a 95% confidence interval and a 5% margin of error. Chi-square tests were utilized to examine associations among dependent variables.

## RESULTS

A flowchart illustrated the study’s outcomes on the knowledge, attitudes, and practices regarding passive smoking among undergraduate students of the University of Abuja, Nigeria. A total of 355 questionnaires were administered for the study, of which 14 were discarded due to errors and incomplete responses, resulting in a response rate of 96.06%. Ultimately, 341 questionnaires were analysed, which equalled the calculated sample size.

Table 1 showed that most respondents were aged 21─25, with 163 respondents (47.8%). The mean age was 21.33±2.6 years. Additionally, there were 189 female respondents (55.4%) compared to 152 males (44.6%). Most respondents identified as Christian, with 281 (82.4%), while 58 (17%) identified as Muslim. Most respondents were Yoruba, totalling 90 (22.6%), while Hausa and Igbo respondents were 9 (7.9%) and 87 (25.5%), respectively. Most respondents were single, comprising 326 (95.6%), while 13 (3.8%) were married. Second-year (200L) students made up a substantial portion of the responses, with 109 (32.0%), and most respondents were from the Faculty of Science, totalling 77 (22.6%).

**Table 1:**
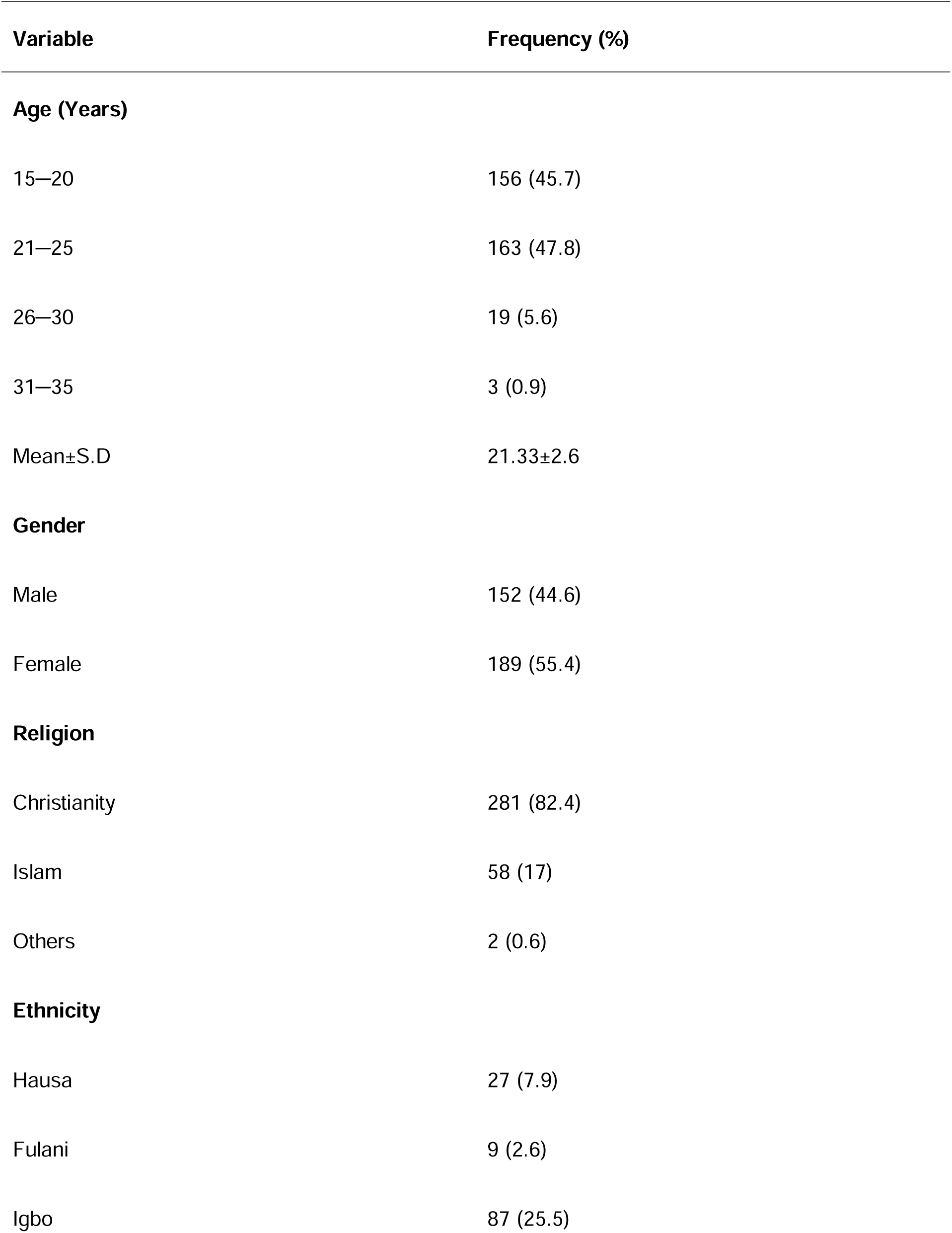
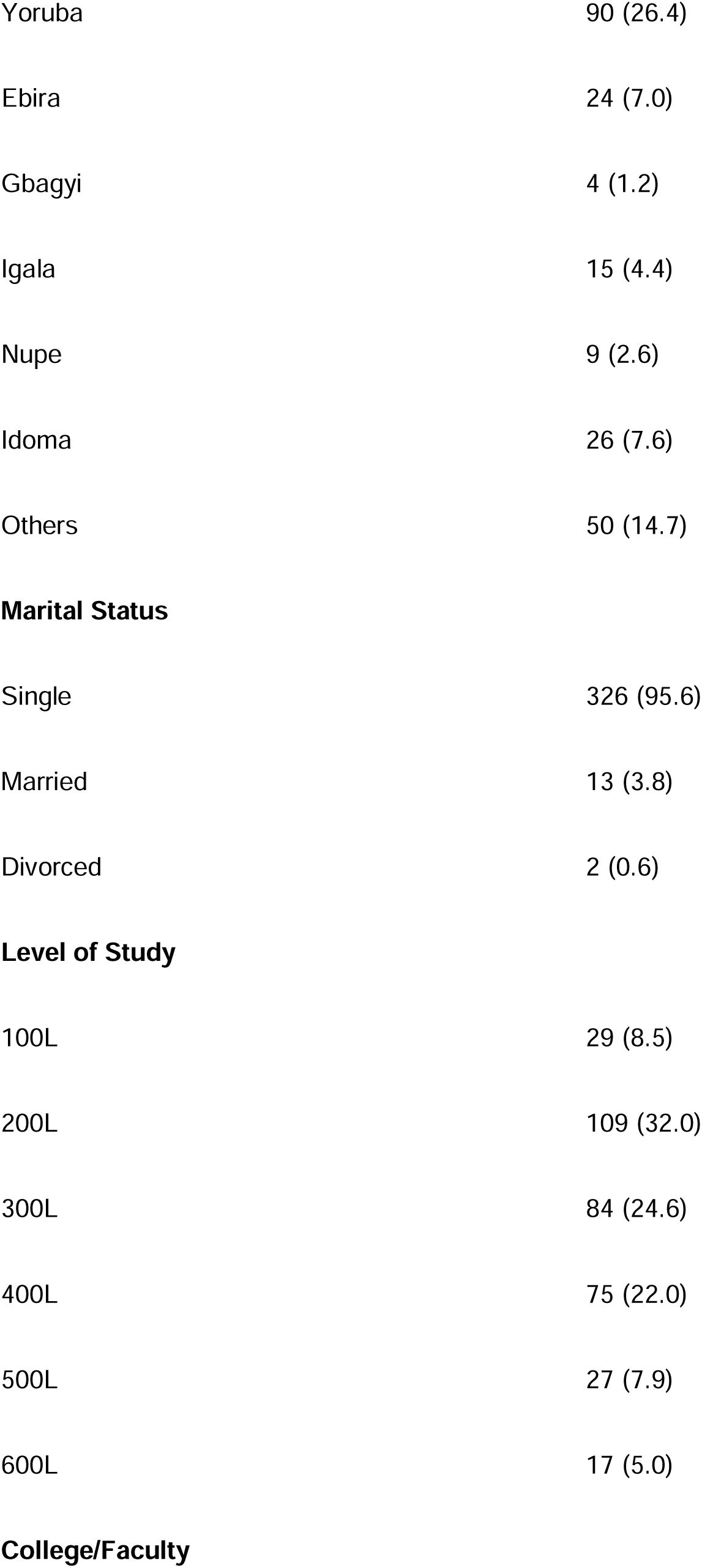
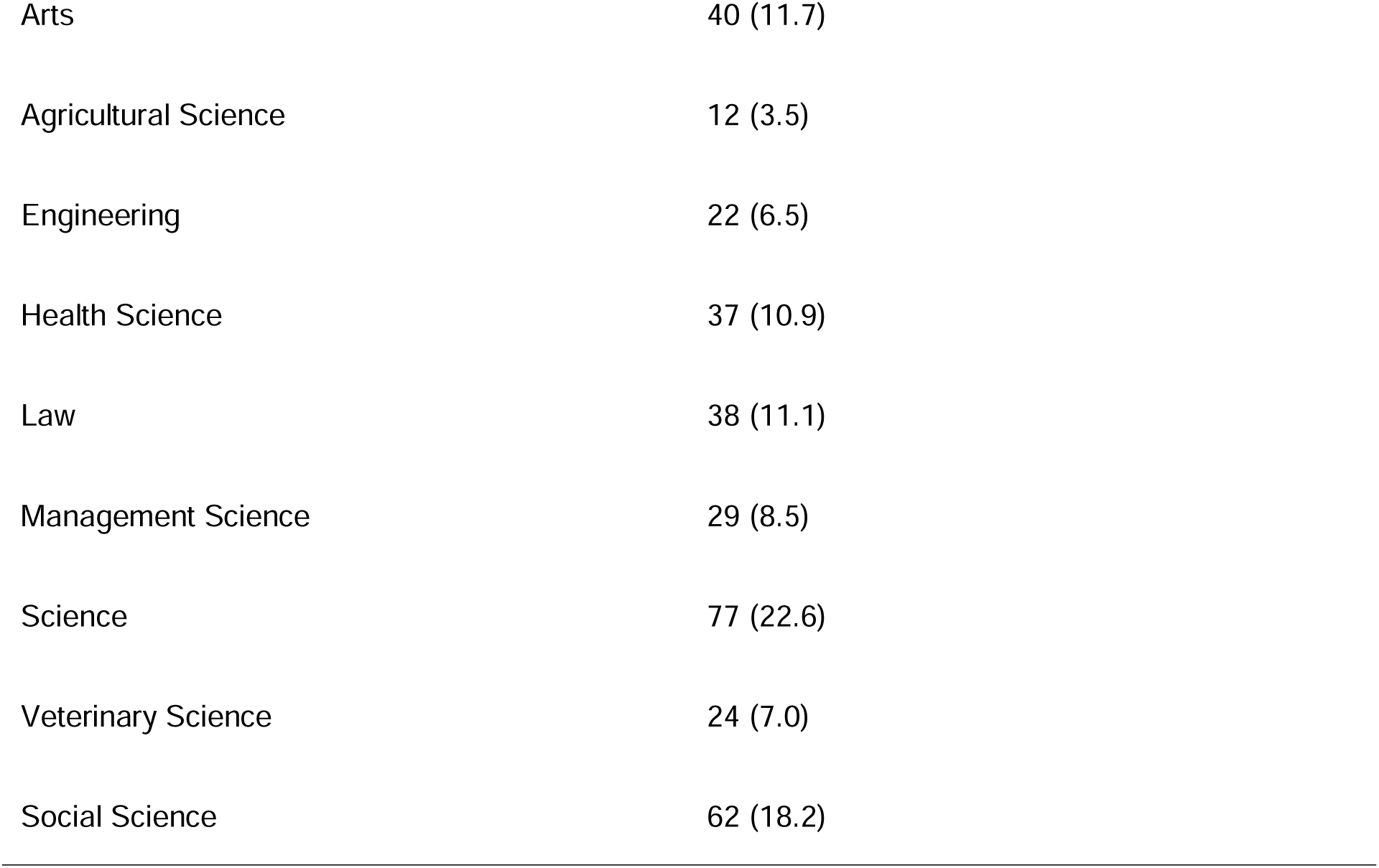
Sociodemographic Characteristics of Respondents.

| **Variable** | **Frequency (%)** |
| --- | --- |
| **Age (Years)** |  |
| 15–20 | 156 (45.7) |
| 21–25 | 163 (47.8) |
| 26–30 | 19 (5.6) |
| 31–35 | 3 (0.9) |
| Mean±S.D | 21.33±2.6 |
| **Gender** |  |
| Male | 152 (44.6) |
| Female | 189 (55.4) |
| **Religion** |  |
| Christianity | 281 (82.4) |
| Islam | 58 (17) |
| Others | 2 (0.6) |
| **Ethnicity** |  |
| Hausa | 27 (7.9) |
| Fulani | 9 (2.6) |
| Igbo | 87 (25.5) |
| Yoruba | 90 (26.4) |
| Ebira | 24 (7.0) |
| Gbagyi | 4 (1.2) |
| Igala | 15 (4.4) |
| Nupe | 9 (2.6) |
| Idoma | 26 (7.6) |
| Others | 50 (14.7) |
| **Marital Status** |  |
| Single | 326 (95.6) |
| Married | 13 (3.8) |
| Divorced | 2 (0.6) |
| **Level of Study** |  |
| 100L | 29 (8.5) |
| 200L | 109 (32.0) |
| 300L | 84 (24.6) |
| 400L | 75 (22.0) |
| 500L | 27 (7.9) |
| 600L | 17 (5.0) |
| **College/Faculty** |  |
| Arts | 40 (11.7) |
| Agricultural Science | 12 (3.5) |
| Engineering | 22 (6.5) |
| Health Science | 37 (10.9) |
| Law | 38 (11.1) |
| Management Science | 29 (8.5) |
| Science | 77 (22.6) |
| Veterinary Science | 24 (7.0) |
| Social Science | 62 (18.2) |

Table 2 showed that the respondents’ knowledge level was analysed using nine questions on knowledge. For each respondent, a score of one was allocated for a ’yes’ response and zero for a ’no’ or ’I don’t know’ answer. For other questions with only one correct answer, as opposed to ’yes,’ ’no,’ and ’I don’t know,’ the correct answer was scored one, while the two incorrect answers were scored zero. A score of five and above denoted good knowledge, while a score below five denoted poor knowledge. The majority of the respondents had good knowledge (80.6%).

**Table 2:**
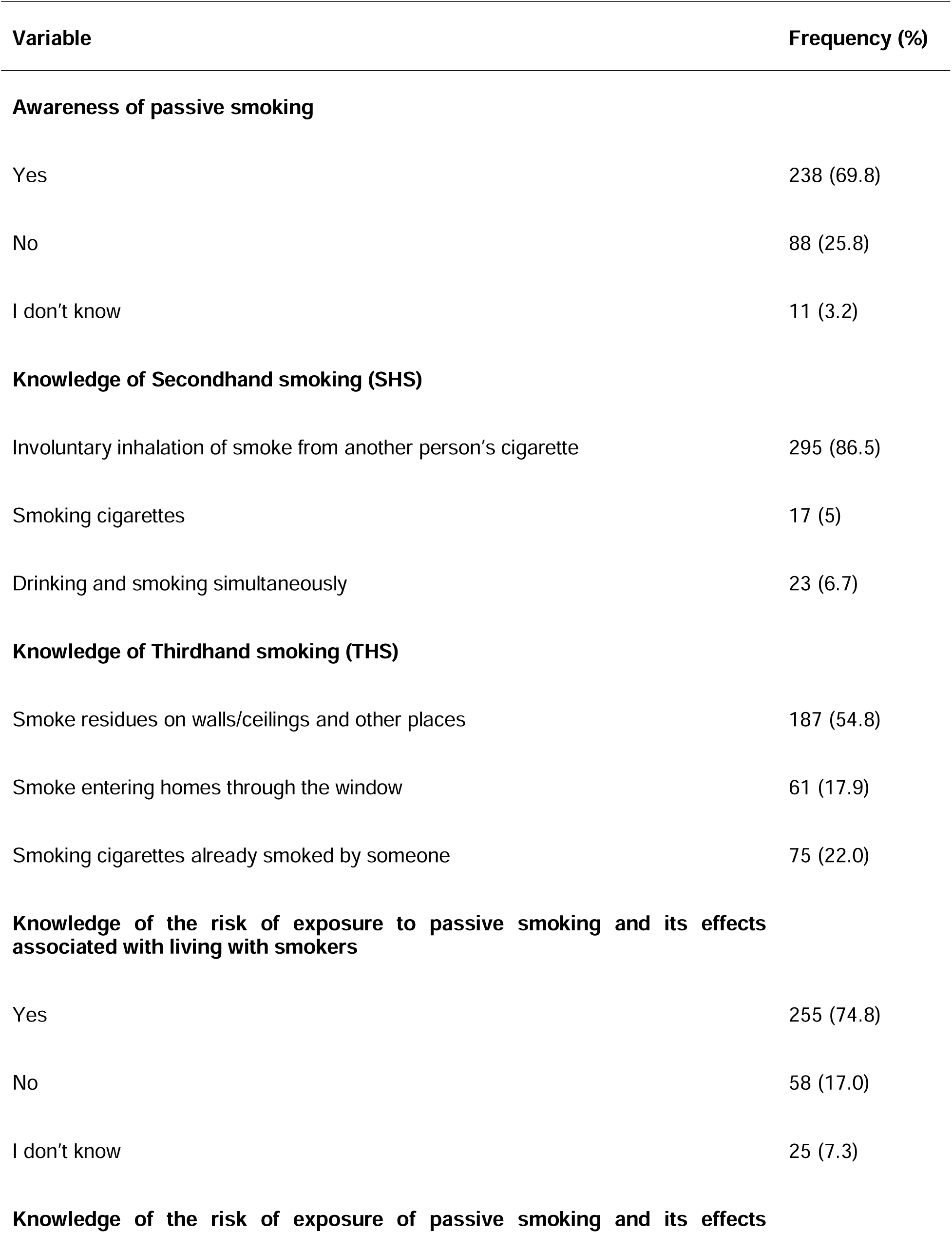
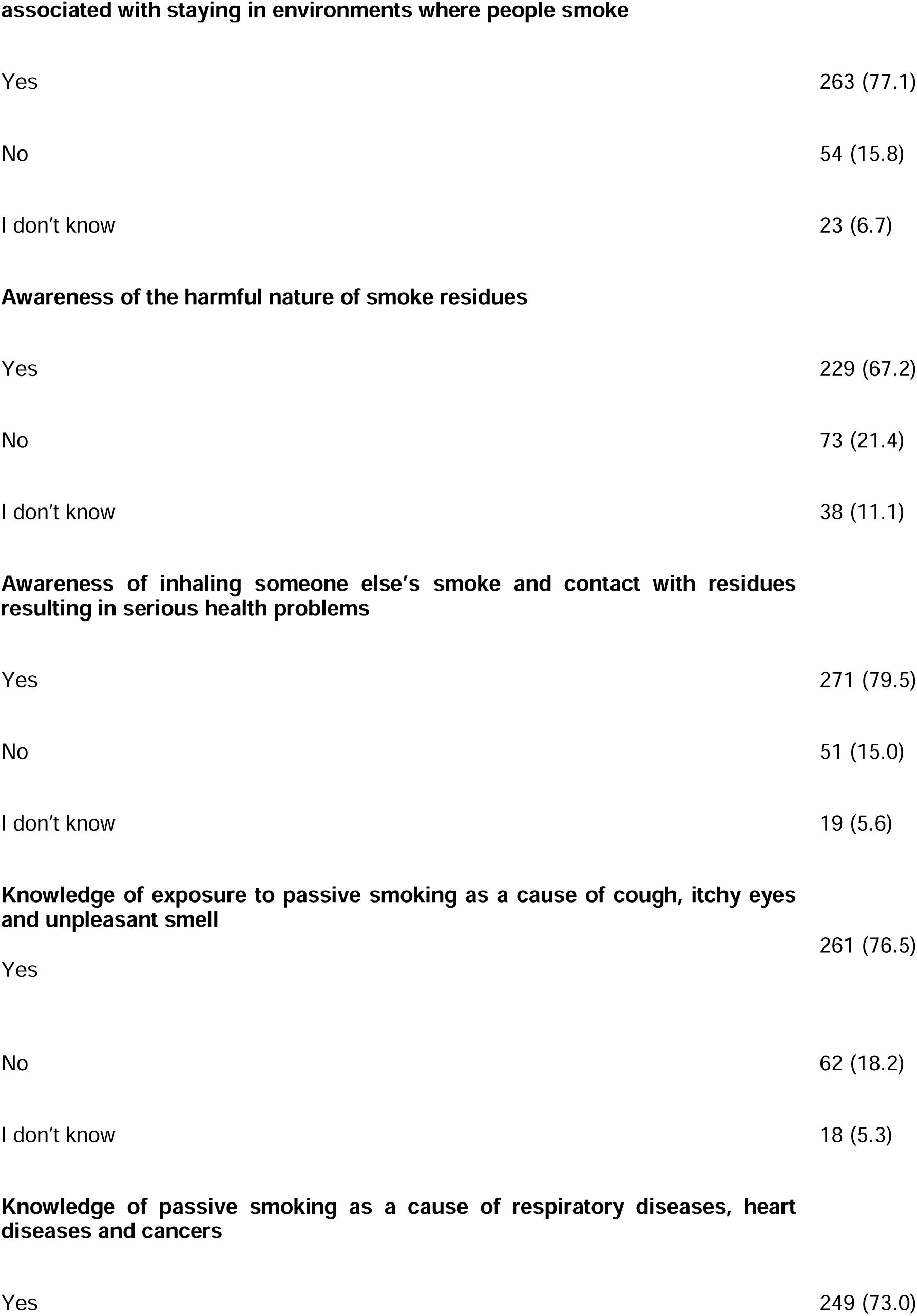
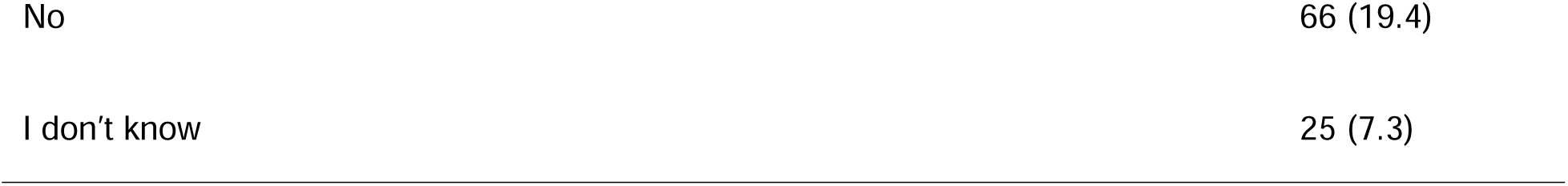
Knowledge of Passive Smoking.

| **Variable** | **Frequency (%)** |
| --- | --- |
| **Awareness of passive smoking** |  |
| Yes | 238 (69.8) |
| No | 88 (25.8) |
| I don't know | 11 (3.2) |
| **Knowledge of Secondhand smoking (SHS)** |  |
| Involuntary inhalation of smoke from another person's cigarette | 295 (86.5) |
| Smoking cigarettes | 17 (5) |
| Drinking and smoking simultaneously | 23 (6.7) |
| **Knowledge of Thirdhand smoking (THS)** |  |
| Smoke residues on walls/ceilings and other places | 187 (54.8) |
| Smoke entering homes through the window | 61 (17.9) |
| Smoking cigarettes already smoked by someone | 75 (22.0) |
| **Knowledge of the risk of exposure to passive smoking and its effects associated with living with smokers** |  |
| Yes | 255 (74.8) |
| No | 58 (17.0) |
| I don't know | 25 (7.3) |
| **Knowledge of the risk of exposure of passive smoking and its effects** |  |
| **associated with staying in environments where people smoke** |  |
| Yes | 263 (77.1) |
| No | 54 (15.8) |
| I don't know | 23 (6.7) |
| **Awareness of the harmful nature of smoke residues** |  |
| Yes | 229 (67.2) |
| No | 73 (21.4) |
| I don't know | 38 (11.1) |
| **Awareness of inhaling someone else's smoke and contact with residues resulting in serious health problems** |  |
| Yes | 271 (79.5) |
| No | 51 (15.0) |
| I don't know | 19 (5.6) |
| **Knowledge of exposure to passive smoking as a cause of cough, itchy eyes and unpleasant smell** |  |
| Yes | 261 (76.5) |
| No | 62 (18.2) |
| I don't know | 18 (5.3) |
| **Knowledge of passive smoking as a cause of respiratory diseases, heart diseases and cancers** |  |
| Yes | 249 (73.0) |
| No | 66 (19.4) |
| I don't know | 25 (7.3) |

Table 3 showed the frequency of positive statements used to assess respondents’ attitudes towards passive smoking, employing a 7-item, 5-point Likert-type question a scale that varied from strongly disagree (score of 1) to strongly agree (score of 5). The mode of the participants for each question was generated, depicting the most frequent responses for each item. The mean of the various items was calculated, and those below the average indicated a poor attitude, while those above the average indicated a good attitude.

**Table 3:** Frequency of Response to Attitude Questions.

| S/N | VARIABLES | SA(5) | A(4) | I(3) | D(2) | SD(1) | MEAN |
| --- | --- | --- | --- | --- | --- | --- | --- |
| 1 | Knowledge of passive smoking and its associated health effects is important to you | 175<br>(875) | 148<br>(592) | 16<br>(48) | 1<br>(2) | 0<br>(0) | 4.45 |
| 2 | People should not smoke where others are because it exposes them to passive smoke | 181<br>(905) | 142<br>(568) | 14<br>(42) | 1<br>(2) | 2<br>(2) | 4.45 |
| 3 | Passive smoking leads to health problems like cancers, heart and respiratory problems | 143<br>(715) | 163<br>(652) | 27<br>(81) | 6<br>(12) | 1<br>(1) | 4.28 |
| 4 | Reducing exposure to passive smoking can be best achieved by developing programs for persons who smoke (such as tobacco free clubs) | 124<br>(620) | 169<br>(676) | 40<br>(120) | 2<br>(4) | 2<br>(2) | 4.17 |
| 5 | Banning smoking in public places can help curtail the excesses of passive smoking | 163<br>(815) | 130<br>(520) | 36<br>(108) | 10<br>(20) | 1<br>(1) | 4.29 |
| 6 | People that violate ban on smoking in public places should be sanctioned | 147<br>(735) | 135<br>(540) | 45<br>(135) | 9<br>(18) | 4<br>(4) | 4.20 |
| 7 | Convincing people around you to quit smoking is a way of preventing passive smoking | 143<br>(715) | 153<br>(612) | 38<br>(114) | 2<br>(4) | 4<br>(4) | 4.25 |

SA = Strongly Agree, A = Agree, I = Indifferent, D = Disagree, SD = Strongly Disagree

Table 4 showed the responses to smoking practices. The responses to the practice questions were scored with values of 1, 2, and 3 for ‘never,’ ‘sometimes,’ and ‘frequently,’ respectively, except for questions 1, 20, 21, 22, and 23. In question 1, a response of ‘yes’ was scored as 3, while ‘no’ and ‘I don’t know’ were each scored as 1. In question 20, the hours of exposure to passive smoke were graded as follows: 0─1 h (1), 2─4 h (2), and 58 h (3). In question 21, a response of ‘yes’ received a score of 3, while ‘no’ and ‘I don’t smoke’ were scored as 1. In questions 22 and 23, scores were assigned to responses as 1, 2, and 3 for ‘frequently,’ ‘sometimes,’ and ‘never,’ respectively. Overall, 55.7% of the respondents reported living with or relating to people who actively smoked, exposing them to passive smoke. Approximately 9.4% believed they were frequently exposed to passive smoke. About 8.5% of the respondents frequently went out to bars to socialize, and around 5.9% spent as much as 5─8 h in those bars. Additionally, 11.1% of the respondents separated themselves from others, and 5.3% frequently disposed of smoke residues to reduce their chances of exposure to passive smoke from those who smoked. Just over 11.4% of the respondents observed that people who smoked around them frequently disposed of their smoke residues, while 13.8% reported coming into contact with smoke residues.

**Table 4:** Practice of Passive Smoking.

| **S/N** | **Passive Smoking Practices** | **Never (%)** | **Sometimes (%)** | **Frequently (%)** |
| --- | --- | --- | --- | --- |
| 1 | Frequency of exposure to passive smoking in offices, hostels, homes or off-campus apartments | Yes (%)<br>190 (55.7%) | No (%)<br>87 (25.5%) | I don't know (%)<br>63 (18.8%) |
| 2 | Frequency of exposure to passive smoking from others smoking | 75 (22%) | 186 (54.5%) | 32 (9.4%) |
| 3 | Frequency of going to bars to socialize | 134 (39.3%) | 175 (51.3) | 29 (8.5%) |
| 4 | Hours spent in bars | 0-1<br>80 (23.5%) | 2-4<br>142 (41.6%) | 5-8<br>20 (5.9%) |
| 5 | Practice of separating oneself from others while smoking to reduce their chances of being exposed to passive smoking | Yes<br>38 (11.1%) | No<br>46 (13.5%) | I don't smoke<br>242 (71.3) |
| 6 | Frequency of disposing residues among those who smoke | 67 (19.6%) | 43 (12.6%) | 18 (5.3%) |
| 7 | Frequency of others disposing their residues among those exposed to passive smoking | 72 (21.1%) | 192 (56.3%) | 39 (11.4%) |
| 8 | Practice of contact with smoke residues | 62 (18.2%) | 219 (64.2%) | 47 (13.8%) |

Table 5 showed varying levels of attitude, knowledge, and practice regarding passive smoking among respondents. Most respondents demonstrated good knowledge, with 275 respondents (80.6%) exhibiting this understanding, while 66 respondents (19.4%) exhibited poor knowledge. Most respondents had a positive attitude towards passive smoking, totalling 337 respondents (98.8%), compared to only 4 (1.3%) with a negative attitude. In terms of practice, 206 respondents (60.4%) reported being exposed to passive smoking, while 135 respondents (39.6%) identified as unexposed.

**Table 5:**
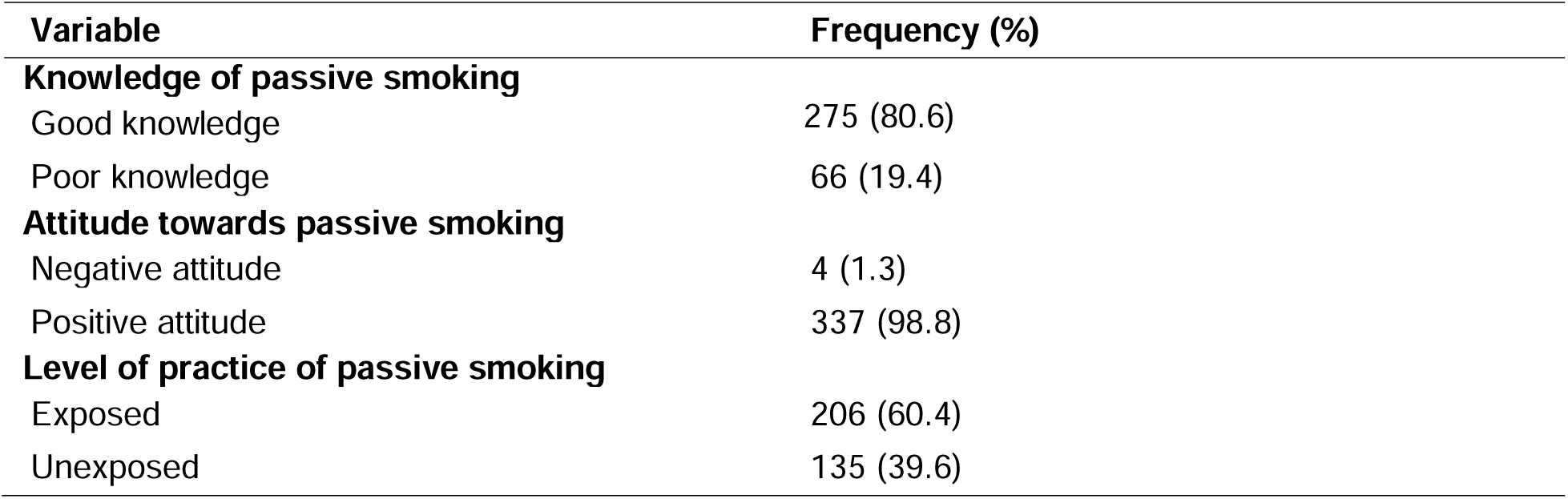
Knowledge, Attitudes, and Practices Regarding Passive Smoking among Respondents.

Table 6 showed the cross-tabulation of knowledge, attitude, and practice regarding passive smoking against age, level of study, and gender, along with the respective chi-square (χ²) values and p-values. Statistically significant relationships were identified between knowledge and the level of study (p < 0.05), indicating that knowledge of passive smoking increased with the level of study. Additionally, a significant association was observed between attitude towards passive smoking and age (p < 0.05), suggesting that attitudes improved with increasing age. No significant correlations were identified between the practice of passive smoking and the assessed demographic variables.

**Table 6:**
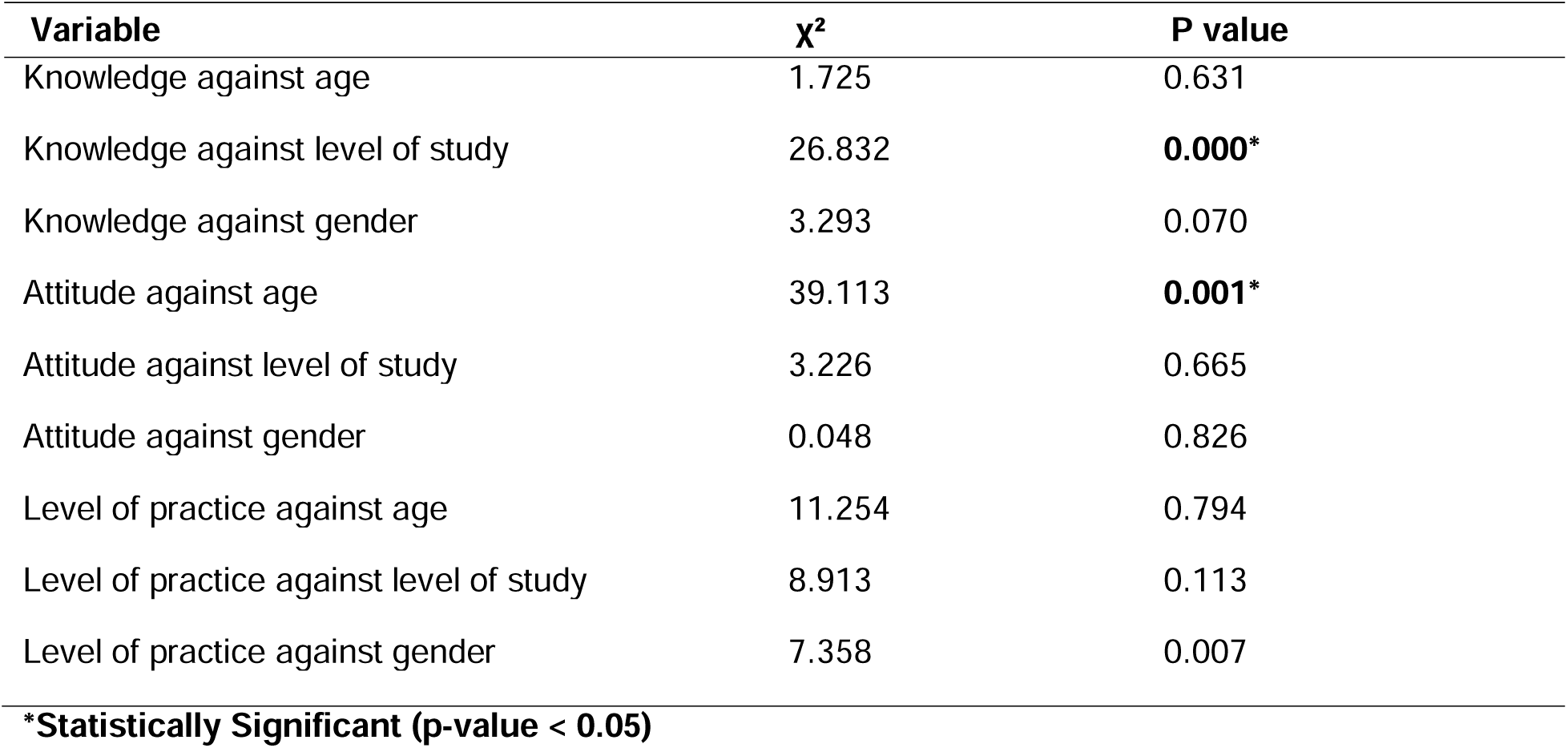
Associations of Age, Level of Study, and Gender with Knowledge, Attitude, and Practice of Passive Smoking.

## DISCUSSION

This study aimed to investigate the attitude, knowledge, and practice of passive smoking among undergraduate students at the University of Abuja. The demographic characteristics of the respondents are presented in Table 1, indicates that the majority of participants were aged 21─25 years, with an average age of 21.33 ± 2.6 years. This is consistent with earlier studies conducted in Eastern Nigeria, where a comparable age distribution was observed among students.^22^ This young demographic is particularly relevant, as younger individuals are often more experimental and may be more susceptible to passive smoke exposure, raising concerns about their health outcomes.

The gender distribution revealed a higher proportion of female respondents (55.4%) compared to males (44.6%). This trend is consistent with findings from other studies,^22^ suggesting that female students may be more willing to participate in such research. Additionally, our findings revealed that most respondents were single, with a significant number of second-year students (32%), which contrasts with previous studies that found varying distributions across different academic years.^22,23^

In terms of knowledge, the study found that 80.6% of respondents demonstrated a solid understanding of passive smoking and its health consequences, as detailed in Table 2. This is consistent with studies conducted among undergraduate students in other countries,^24,25^ indicating a global trend of heightened awareness among students regarding the dangers of secondhand smoke. Such awareness may stem from substantial public health campaigns promoting smoking cessation and educating the public about the risks associated with both smoking and passive exposure, particularly in developed countries

The respondents exhibited overwhelmingly positive attitudes, with 98.8% expressing a desire to avoid passive smoke exposure (Table 3). This indicates a promising trend in student perceptions toward tobacco control. However, it is essential to highlight that positive attitudes do not compulsorily translate into behavioural changes, as evidenced by the 60.4% prevalence of reported passive smoke exposure (Table 5). Similar findings were reported in a study among women at two universities in Jordan.^24^ However, this contrasts with another study where the prevalence of passive smoke exposure was reported to be 38.8% among non-smoking adults in Nigeria. Additionally, it differs from a research study carried out with medical students in Turkey.^25^ The discrepancy between knowledge and practice reflects a common gap observed in tobacco control literature, where a majority of respondents agreed that tobacco smoke poses a threat to the health of non-smokers and contributes to heart disease, triggers asthma attacks, and can lead to lung cancer.^24, 26, 27^ This emphasizes the need for targeted behavioural interventions. The significant correlation between knowledge and academic level, highlighted in Table 6, underscores the value of educational interventions as students’ progress in their studies. Our findings suggest that as students advance academically, their understanding of the adverse effects of passive smoking increases.

The high exposure rate to passive smoke among respondents raises concerns about the effectiveness of existing tobacco control measures in Nigeria. Despite legislation aimed at regulating the consumption of tobacco and exposure to tobacco smoke, as outlined in the WHO FCTC,^28^ our findings suggest that implementation remains inadequate, contributing to continued passive smoke exposure among university students. This is particularly alarming given the well-documented health risks associated with secondhand smoke, including respiratory diseases and cardiovascular conditions.

Our study’s limitations include potential recall bias, as the data relied on self-reported questionnaires, which may lead to inaccuracies in reporting exposure levels. Furthermore, social desirability bias may have influenced respondents to provide answers they deemed more socially acceptable, rather than their true experiences. These limitations must be acknowledged, as they may affect the reliability of the results.

The generalizability of these findings may extend beyond the University of Abuja, offering insights applicable to other universities and similar educational settings in Nigeria and other developing countries. However, variations in local cultural practices, regulatory environments, and public health initiatives must be considered when extrapolating these results to broader populations.

In conclusion, while the students at the University of Abuja exhibit a commendable level of knowledge and a positive attitude toward passive smoking prevention, the high prevalence of exposure indicates a significant gap between awareness and practice. To address this, effective public health strategies are essential. These should include comprehensive tobacco control policies, educational campaigns, and strict enforcement of smoke-free regulations in public spaces. Collaborative initiatives between university authorities and student organizations, such as the Tobacco Free Club (TFC-Uniabuja), can further enhance awareness and participation in tobacco prevention efforts. These measures are vital for safeguarding the health of non-smokers and advancing a smoke-free future in Nigeria.

## Author’s contribution

Conceptualization: G.K.J, S.I.A, I.I, P.T.A, M.A.O. Data Curation: G.K.J, S.I.A, I.I, P.T.A, M.A.O. Formal Analysis: G.K.J, S.I.A, I.I, P.TA., M.A.O. Funding Acquisition: G.K.J, S.I.A, I.I, P.T.A, M.A.O. Investigation: G.K.J, S.I.A, I.I, P.T.A, M.A.O. Methodology: G.K.J, S.I.A, I.I, P.T.A M.A.O. Resources: G.K.J, S.I.A, I.I. Software: G.K.J. Supervision: G.K.J, S.I.A. Validation: G.K.J, S.I.A, I.I, M.A.O. Visualization: G.K.J, S.I.A, I.I, P.T.A, M.A.O. Writing - Original Draft: G.K.J, S.I.A, I.I, P.T.A, M.A.O. Writing - Review Editing: G.K.J, S.I.A, I.I, P.T.A, M.A.O.

## Acknowledgements

We are indebted to the entire Community Medicine Division at the University of Abuja. We are especially grateful to Dr. Mustapha Abubakar Jamda, a Community Medicine and Public Health professor, for supervising this project and for the role of mentorship he played in it.

## Data Availability Statement

All data produced in the present study are available upon reasonable request to the authors.

## References

1. Bristol Public Health. Health and wellbeing factsheet. Available from https://www.bristol.gov.uk/council/policies-plans-and-strategies/social-care-and-health/joint-strategic-needs-assessment/health-and-wellbeing-reports. cited October 21, 2024

2. Desalu O, Onyedum C, Adewole O, Fawibe E and Salami, K. Secondhand smoke exposure among non-smoking adults in two Nigeria cities. Ann Afr Med. 2011;10:103–111.

3. Adepoju E, Olowookere S, Adeleke A, Afolabi O, Olajide F, Aluko O. A population-based study on the prevalence of cigarette smoking and smokers’ characteristics at Oshogbo, Nigeria. Tobacco Use Insights. 2013;6:1–5.

4. Hopkinson S, Lester-George A, Ormiston-Smith N, Cox A, Arnott D. Child uptake of smoking by area across the UK. Thorax. 2013;69(9).

5. Jallow IK, Britton J, Langley T. Prevalence and factors associated with exposure to second hand smoke (SHS) among young people: a cross-sectional study from the Gambia. BMJ Open. 2017;8(3).

6. Jiang XQ, Mei XD, Feng D. Air pollution and chronic airway diseases: what should people know and do? J Thorac Dis. 2016;8(1):E31–40.

7. World Health Organization. Global estimate of the burden of disease from second hand smoke. 2010. Available from: https://www.who.int/publications/i/item/9789241564076. Last updated July 30, 2010; cited October 21, 2024

8. World Health Organization. Tobacco. WHO health topics. 2021. Available from https://www.who.int/health-topics/tobacco#tab=tab_3. cited October 21, 2024.

9. Udi OA. Student nurses exposure to passive smoking at a community in Eastern Nigeria. Int J Res. 2019;06(04).

10. Northrup TF, Jacob III P, Benowitz NL, Hoh E, Quintana PJE, Hovell MF, Matt GE, Stotts AL. Thirdhand smoke: state of the science and a call for policy expansion. Public Health Rep. 2016;131(2):233–238.

11. Al-Kubaisy W, Abdullah NN, Al-Nuaimy H, Kahn S M, Halawany G, Kurdy S. Factors associated with smoking behaviour among university students in Syria. Procedia. 2012;38:59–65.

12. Nnodu OE, Jamda MA. Exposure of undergraduate students to cigarette adverts: A case study of University of Abuja, Nigeria. J Med Trop 2014;16(2):93–96.

13. Perez DA, Grunseit AC, Rissel C, Kite J, Cotter T, Dunlop S, et al. Tobacco promotion ’below-the-line’: exposure among adolescents and young adults in NSW, Australia. BMC Public Health. 2012;12:429.

14. Alanazi A, Al Enezi F, Alqahtani MM, Alshammari TF, Ansari MA, Al-Oraibi S, et al. Effects of passive smoking on students at College of Applied Medical Sciences, King Saud bin Abdulaziz University for Health Sciences. 2015;6(1):100–105.

15. Golden RN, Fiore MC. The 50th anniversary of the Surgeon General’s Report on Smoking and Health: reflections and lessons to be learned for other public health challenges. WMJ. 2014. 1;113(2):81–2.

16. Harvim P, Zhang H, Georgescu P. Cigarette smoking on college campuses: an epidemiological approach. J. Appl. Math. Comput. 2021;65:515–540.

17. Al-Kubaisy W, Abdullah NN, Al-Nuaimy H, Kahn S M, Halawany G, Kurdy S. Factors associated with smoking behaviour among university students in Syria. Procedia. 2012;38:59–65.

18. Perez DA, Grunseit AC, Rissel C, Kite J, Cotter T, Dunlop S, et al. Tobacco promotion ’below-the-line’: exposure among adolescents and young adults in NSW, Australia. BMC Public Health. 2012;12:429.

19. Tobacco Control Laws. Legislation by country: Nigeria. Available from https://www.tobaccocontrollaws.org/legislation/country/nigeria/summary. Last updated July 26, 2021; cited October 21, 2024.

20. Vu GV, Ngo CQ, Phan PT, Doan LPT, Nguyen TT, Nguyen MH et al. Inadequate knowledge, attitude and practices about second-hand smoke among non-smoking pregnant women in urban Vietnam: the need for health literacy reinforcement. Int J Environ Res Public Health. 2020;17(10):3744.

21. Haddad C, Sacre H, Hajj A. Comparing cigarette smoking knowledge and attitudes among smokers and non-smokers. Environ Sci Pollut Res. 2020;27:19352–19362.

22. Udi OA. Student Nurses Exposure to Passive Smoking at a Community in Eastern Nigeria. Int J Res. 2019;6(4). Available from: https://journals.pen2print.org/index.php/ijr/.

23. Juraybi A, Arishy A, Qussairy E, Majrashi E, Alfaifi K, Al-Musalam J, Dyab OA, Yassin A. Awareness about passive smoking among Jazan University students, Saudi Arabia Med Sci. 2021:25(110):973–984.

24. Gharaibeh H, Haddad L, Alzyoud S, El-Shahawy O, Abu-Baker N, Umlauf M. Knowledge, attitudes, and behavior in avoiding secondhand smoke exposure among non-smoking employed women with higher education in Jordan. Int J Environ Res Public health. 2011;8(11):4207–4219

25. Desalu O, Onyedum C, Adewole O, Fawibe E and Salami, K. Secondhand smoke exposure among non-smoking adults in two Nigeria cities. Ann Afr Med. 2011;10:103–111

26. Mishra S, Thind H, Gokarakonda S, Lartey G, Watkins C, Chahal, M. Second-hand smoke in a University campus: Attitudes and perceptions of faculty, staff and students. 2011;4(1):21–27.

27. King B, Dube S, Babb S. Perception about the harm of secondhand smoke exposure among U.S middle and high school students: findings from the 2012 National Youth Tobacco Survey. Tobacco-induced diseases 2013;11(16):1–5.

28. World Health Organization. (2015). The WHO framework convention on tobacco control: 10 years of implementation in the African region. Available from: https://apps.who.int/iris/bitstream/handle/10665/164353/9789290232773_eng.pdf. cited Oct 21, 2024

